# mTOR Inhibitor-Based Immunosuppression Is Associated with Lower Parathyroid Hormone Levels in Kidney Transplant Recipients: A Multinational Database Analysis and Longitudinal Single-Center Study

**DOI:** 10.64898/2026.05.16.26353370

**Authors:** Yael Lein, Iddo Z. Ben-Dov, Keren Tzukert

## Abstract

Secondary hyperparathyroidism persists in the majority of kidney transplant recipients and is associated with adverse graft and cardiovascular outcomes. The immunosuppressive drug class used post-transplant may modulate parathyroid hormone (PTH) levels through distinct mechanisms: calcineurin inhibitors (CNI) stabilize PTH mRNA, while mTOR inhibitors (mTORi) suppress parathyroid cell proliferation in experimental models. We report supporting evidence from two independent analyses. In a multinational real-world database analysis (TriNetX Global Collaborative Network), kidney transplant recipients with documented mTORi use and eGFR in the target range had lower PTH than those on CNI across eGFR strata examined (15-30, 30-45, 45-60, 60-75, >75 mL/min/1.73 m^2^), with risk ratios for PTH >130 pg/mL ranging from 0.47 to 0.67 in propensity-matched analyses (all p < 0.05). The known confounders - calcium (higher in CNI) and phosphate (higher in mTORi) - both act to oppose this pattern, strengthening the possibility of a drug effect. In a longitudinal single-center cohort (n = 118; 796 PTH measurements), a linear mixed-effects model with time-varying mTORi exposure confirmed a 42% lower PTH during on-mTORi periods after adjustment for eGFR, transplant vintage, diabetes, age, and sex (fold-change 0.58 [95% CI 0.50-0.68]; p < 0.0001). These findings suggest a direct PTH-lowering effect of mTORi. Immunosuppression choice may be considered in the management of post-transplant hyperparathyroidism in selected patients.

## Introduction

Secondary hyperparathyroidism is a common disorder following kidney transplantation, persisting in 50-70% of recipients beyond the first year and independently predicting graft failure, cardiovascular events, and mortality [1]. While improved renal phosphate excretion and correction of acidosis after transplantation partially normalizes PTH, hypercalcemia, inadequate calcitriol synthesis, and autonomous parathyroid gland function sustain elevated PTH in many patients [2,3].

The choice of maintenance immunosuppression is determined primarily by rejection risk and tolerability [4,5], but may have underappreciated effects on mineral metabolism. CNI - the backbone of most immunosuppression regimens - have been shown to stabilize PTH mRNA through post-transcriptional mechanisms, augmenting PTH secretion independently of calcium or phosphate signaling [6]. Conversely, mTOR inhibitors (sirolimus, everolimus) suppress parathyroid cell proliferation in rodent models of secondary hyperparathyroidism [7], raising the hypothesis that mTORi-based immunosuppression may attenuate post-transplant hyperparathyroidism.

However, mTORi recipients tend to have better graft function and lower phosphate levels - both of which independently suppress PTH. Unraveling a drug effect despite these metabolic confounders requires either within-patient longitudinal designs or very large sample sizes. We attempted both.

## Methods

We used the TriNetX Global Collaborative Network (191.6 million patients; 178 participating healthcare organizations; data refresh 14 May 2026) [8] to identify adult kidney transplant recipients stratified by immunosuppression: Cohort 1 (mTORi, defined by ICD code Z79.623) versus Cohort 2 (CNI: tacrolimus or cyclosporine, without concurrent mTORi). Cohorts were further stratified into five eGFR strata (15-30, 30-45, 45-60, 60-75, >75 mL/min/1.73 m^2^) using MDRD eGFR, yielding ten independent comparisons. Cohort entry (index event) was defined as the most recent transplant procedure, with the requirement that an eGFR measurement in the target range and documentation of the qualifying immunosuppressive regimen (mTORi for Cohort 1; CNI without concurrent mTORi for Cohort 2) both occurred on or after that transplant. The eGFR measurement used for stratum assignment was the same measurement that fulfilled the cohort entry criterion, not a separate assessment. Laboratory outcomes were assessed in the window from 1 day to 1 year after the index event. For continuous laboratory values (PTH, calcium, phosphate), the most recent measurement within this window was used. According to TriNetX outcome reporting conventions, for the PTH >130 pg/mL outcome, a patient was considered positive if any measurement in this window exceeded the threshold. For each stratum, both unmatched and propensity score-matched (1:1) analyses were performed, matching for age, sex, race and diabetes. Outcomes were: (1) PTH levels; (2) proportion with PTH >130 pg/mL (□ 2× upper limit of normal). Potential confounders - calcium and phosphate - were extracted in parralel. However, in its basic functionality, the platform does not provide indivual patient data and thus prohibiting correlation analysis and multivariable adjustment.

We thus conducted a retrospective cohort study of 121 kidney transplant recipients at Hadassah Medical Center, Jerusalem. The Helsinki Committee of the Hadassah Medical Organization gave ethical aproval for this work. Sixty-one patients were exposed to mTORi (36 as CNI+mTORi addition, 25 as CNI to mTORi switch) and 60 received CNI only. mTORi exposure was modeled as a time-varying covariate. Of 121 enrolled patients, 118 contributed at least one PTH measurement and were included in the analyses. All available serial PTH measurements (n = 796 from 118 patients) were log-transformed and analyzed using a linear mixed-effects model (LMM) with a random intercept per patient, adjusting for concurrent eGFR (CKD-EPI), transplant vintage, diabetes, age, and sex.

All statistical analyses and graphical presentations were conducted on R [9], using the packages lme4 and lmerTest for linear mixed-effects modelling, and ggplot2, patchwork, and ggtext for data visualization. We used artificial inteligence (Claude, Anthropics) to assist with R coding, which was executed independetnly by the authors.

## Results

Across all five eGFR strata in the TriNetX analysis, mTORi recipients had lower mean post-index PTH than CNI recipients (Figure 1). In unmatched analyses, mean PTH ranged from 123-157 pg/mL in mTORi versus 146-186 pg/mL in CNI, with statistically significant differences in three of five strata (eGFR 30-45, 45-60, and 60-75). In propensity-matched 1:1 analyses, the direction was consistent across all strata, though p-values were attenuated due to reduced sample sizes. The risk ratio for PTH >130 pg/mL significantly favored mTORi in all five matched strata (RR 0.47-0.67; all p < 0.05) and all five unmatched strata (RR 0.40-0.50; all p < 0.001), with the largest effect at lower eGFR (Figure 2).

**Figure 1.**
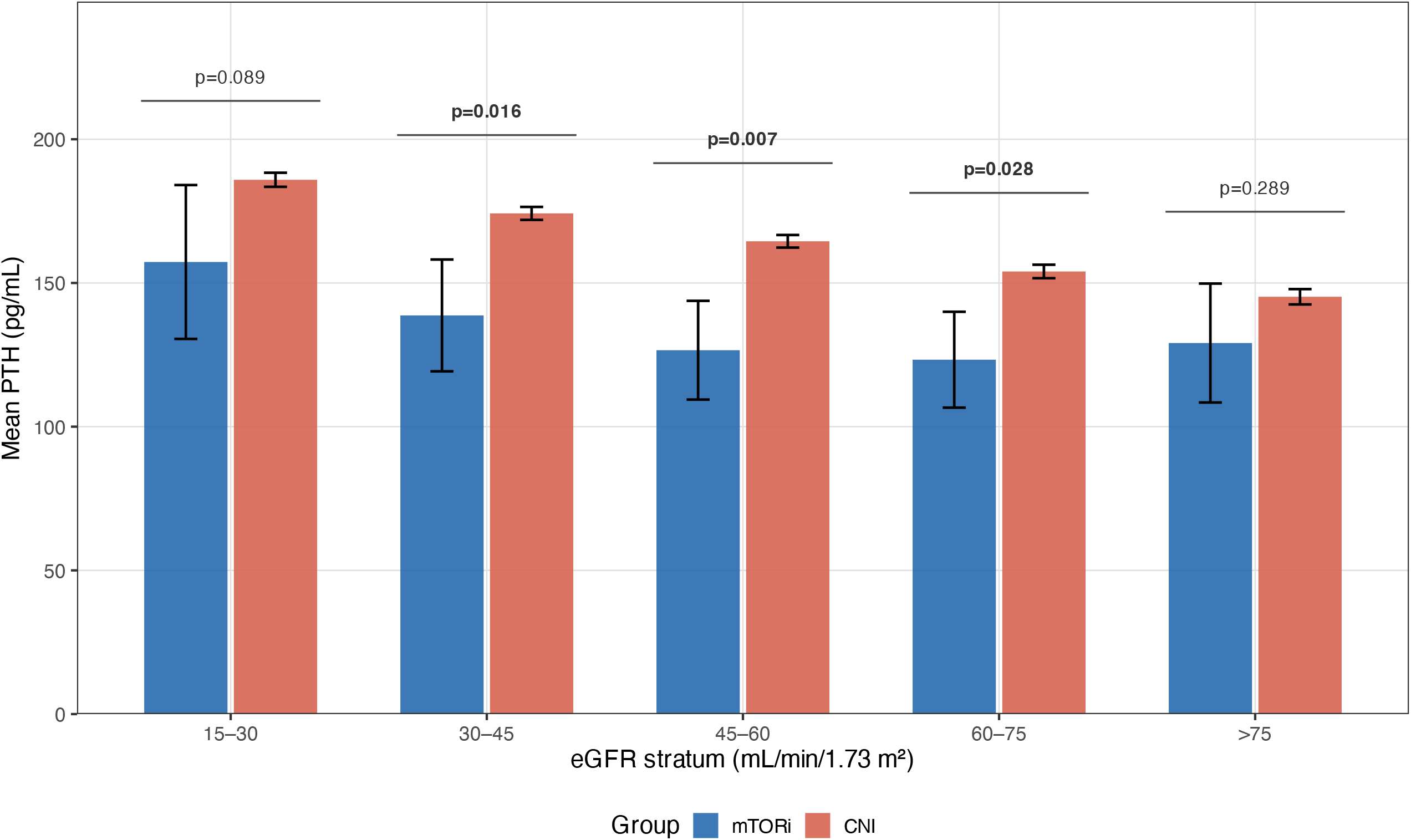
Mean post-index parathyroid hormone (PTH) level by treatment group and eGFR stratum. Bars show mean PTH (pg/mL) with 95% confidence intervals for kidney transplant recipients on mammalian target of rapamycin inhibitors (mTORi, blue) versus calcineurin inhibitors (CNI, red-orange), stratified by estimated glomerular filtration rate (eGFR, mL/min/1.73 m^2^). Analyses are unmatched (full real-world sample). Horizontal brackets indicate between-group comparisons; p-values are from Welch’s t-test (bold = p<0.05). mTORi group sizes: n=121-178 per stratum; CNI group sizes: n=14,929-49,761 per stratum.

**Figure 2.**
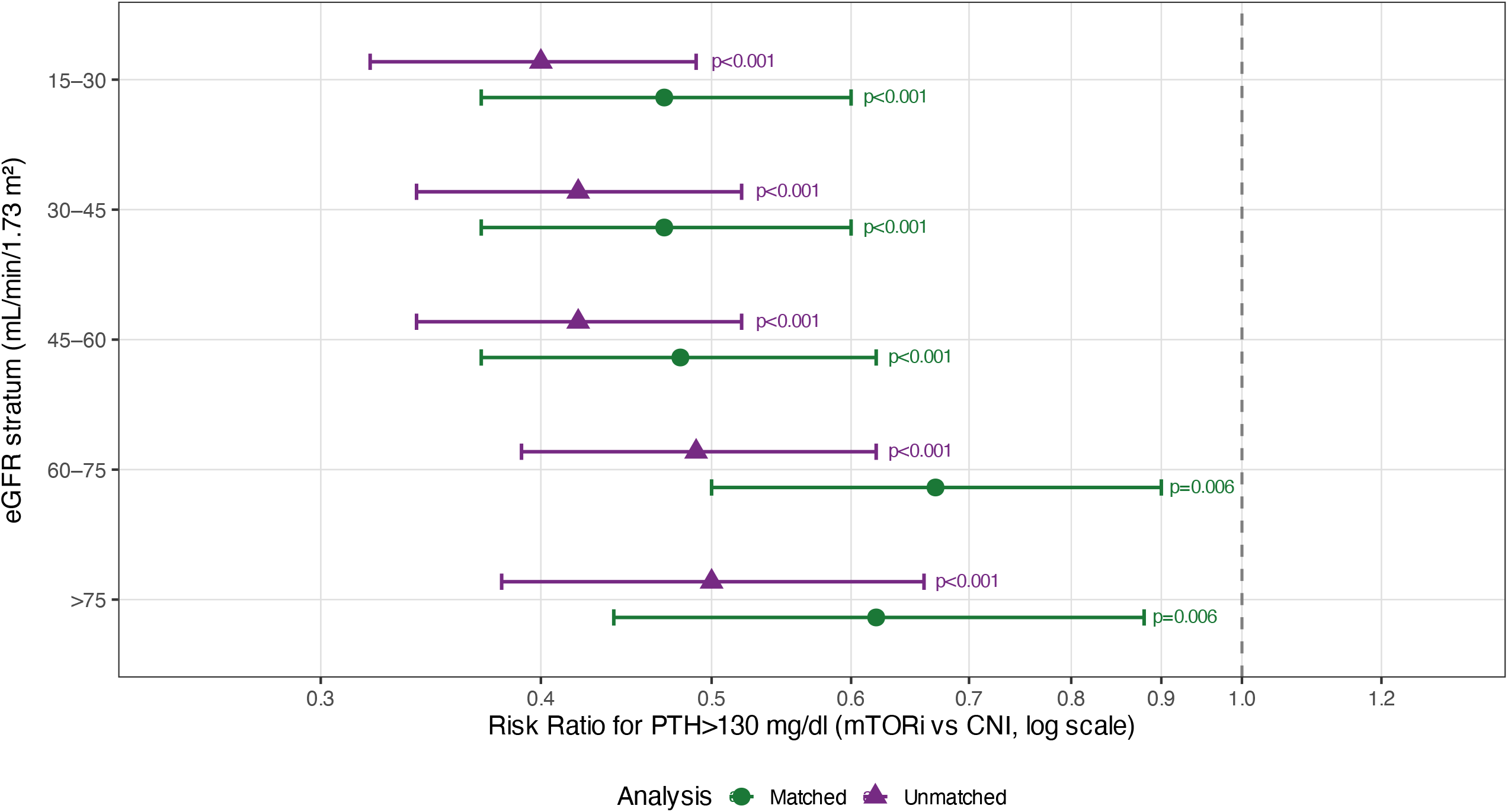
Risk ratios for PTH exceeding 130 pg/mL in mTORi versus CNI recipients, by eGFR stratum. Each point represents the proportion-based risk ratio (RR) with 95% confidence interval, shown separately for propensity score-matched (circles, green) and unmatched (triangles, purple) analyses. RR<1 indicates a lower proportion of mTORi recipients with at least one PTH measurement exceeding 130 pg/mL at any point during the 1-day to 1-year post-index window compared to CNI recipients. p-values are shown to the right of each confidence interval. *Both figures use data from the TriNetX Global Collaborative Network (data download 14 May 2026; total network n=191,649,352). Cohort 1 = mTORi; Cohort 2 = CNI*.

Examination of potential confounders revealed that calcium levels were higher in CNI-versus mTORi-treated patients across most strata (difference 0.21-0.28 mg/dL; p < 0.0001 in 4/5 strata), while phosphate was higher in mTORi versus CNI (difference 0.15-0.33 mg/dL; p < 0.05 in 4/5 strata) (Supplementary Figure 1). Results extracted from the TriNetX Compare Outcomes analyses are summarized in Supplementary Tables 1-3.

In out single-center longitudinal cohort, the dataset comprised 796 PTH measurements from 118 patients. In the LMM, intraclass corrlation coefficient (ICC) was 0.39, indicating substantial within-patient correlation ot PTH levels. On-mTORi periods were associated with 42% lower PTH compared to off-mTORi periods in the same patients after full covariate adjustment (fold-change 0.58 [95% CI 0.50-0.68]; p < 0.0001). Both exposure types showed significant independent effects: CNI+mTORi addition (0.49× [0.41-0.60]; p < 0.0001) and CNI to mTORi switch (0.69× [0.56-0.86]; p = 0.001). Transplant vintage was an independent predictor of lower PTH (p < 0.0001); eGFR, sex and diabetes were *not* independently associated with PTH after adjustment, and neither did age, lower calcium and higher phosphate concentrations (that showed significane in univariable models). Figure 3 shows adjusted PTH values over time (transplantation vintage up to 20 years).

**Figure 3.**
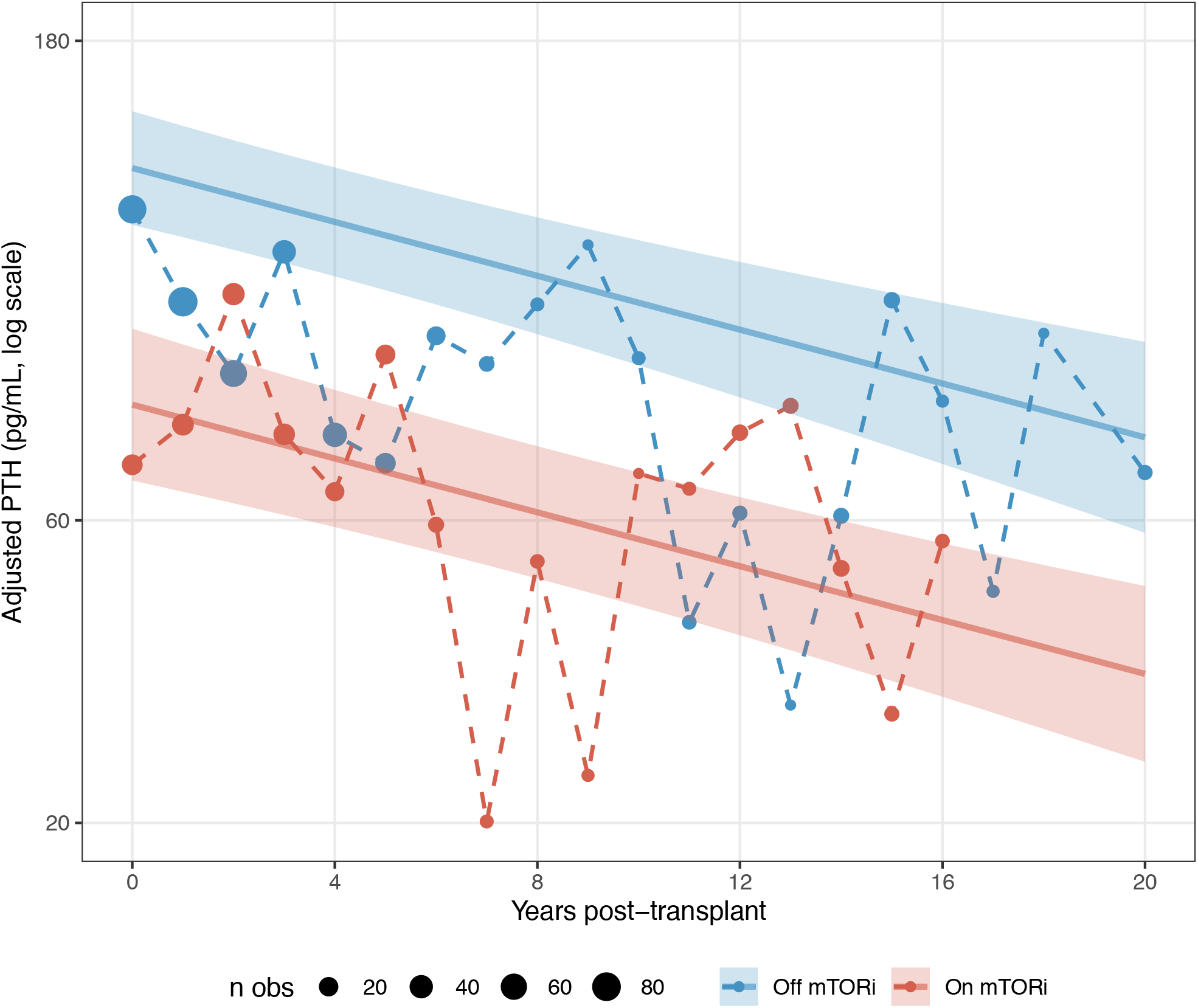
Adjusted PTH by mTORi exposure over transplantation vintage. Dashed lines: observed bin (yearly) medians. Solid lines: Marginal means (95% CI shaded) derived from the following LMM: ‘log(PTH) ∼ on_mTORi + eGFR + vintage + DM + age + sex + (1|patient)’.

## Discussion

This study provides evidence that mTORi-based immunosuppression is associated with lower PTH in kidney transplant recipients, independent of renal function and other measured confounders. The convergence of a large-scale real-world database analysis with a longitudinal cohort design, following the findings in model systems, strengthens causal presumption considerably.

In the global network study, the metabolic differences between mTORi and CNI recipients - higher calcium and lower phosphate in CNI - would each independently suppress PTH in CNI patients relative to mTORi patients. The fact that PTH is nonetheless lower in mTORi recipients, consistently across five eGFR strata and in both matched and unmatched analyses, implies a pharmacological PTH-lowering effect that overcomes these metabolic confounders.

The within-patient design of the single-center LMM eliminates all time-invariant confounders, leaving only time-varying factors (eGFR trajectory, transplant vintage, incident diabetes) which were explicitly adjusted. The 42% PTH reduction on mTORi may be clinically meaningful given the association between elevated post-transplant PTH and graft loss.

The biological mechanism most consistent with these findings is direct mTORi-mediated suppression of parathyroid gland proliferation and PTH synthesis, as demonstrated in rodent models of secondary hyperparathyroidism. The apparent greater effect of CNI+mTORi addition versus CNI to mTORi switch cannot be explained by additive pharmacodynamic action, as CNI were shown in laboratory models to increase PTH secretion.

Limitations include the observational nature of both analyses and the inability to perform multivariable assessments in the TriNetX analysis. The single-center cohort lacks adjustment for additional time-varying confounders such as vitamin D levels and concomitant medications, and is modest in size, though the within-patient LMM design is well-powered for detecting the observed effect magnitude.

## Conclusion

mTOR inhibitor-based immunosuppression is independently associated with lower PTH levels in kidney transplant recipients across a wide range of renal function, as demonstrated in both a large multinational real-world database and a longitudinal single-center cohort. These findings support a direct pharmacological PTH-lowering effect of mTORi and suggest that immunosuppression selection should be considered an active lever in the management of post-transplant secondary hyperparathyroidism.

## Data Availability

All data produced in the present study are available upon reasonable request to the authors

## Figure legends

**Supplementary Figure 1.** Post-Index Calcium and Phosphate Differences. Points = mean difference (mTORi − CNI) ± 95% CI. Both Ca (lower in mTORi) and P (higher in mTORi) would be expected to raise PTH in mTORi relative to CNI. TriNetX Global Collaborative Network, propensity-matched 1:1 analyses.

## References

1. Okada M, Tominaga Y, Sato T, Tomosugi T, Futamura K, Hiramitsu T, Ichimori T, Goto N, Narumi S, Kobayashi T, Uchida K, Watarai Y. Elevated parathyroid hormone one year after kidney transplantation is an independent risk factor for graft loss even without hypercalcemia. BMC Nephrol. 2022 Jun 17;23(1):212. doi: 10.1186/s12882-022-02840-5. PMID: 35710357; PMCID: PMC9205154

2. Evenepoel P, Meijers BK, de Jonge H, Naesens M, Bammens B, Claes K, Kuypers D, Vanrenterghem Y. Recovery of hyperphosphatoninism and renal phosphorus wasting one year after successful renal transplantation. Clin J Am Soc Nephrol. 2008 Nov;3(6):1829–36. doi: 10.2215/CJN.01310308. Epub 2008 Oct 15. PMID: 18922992; PMCID: PMC2572285.

3. Messa P, Sindici C, Cannella G, Miotti V, Risaliti A, Gropuzzo M, Di Loreto PL, Bresadola F, Mioni G. Persistent secondary hyperparathyroidism after renal transplantation. Kidney Int. 1998 Nov;54(5):1704–13. doi: 10.1046/j.1523-1755.1998.00142.x. PMID: 9844148.

4. Halloran PF. Immunosuppressive drugs for kidney transplantation. N Engl J Med. 2004 Dec 23;351(26):2715–29. doi: 10.1056/NEJMra033540. Erratum in: N Engl J Med. 2005 Mar 10;352(10):1056. PMID: 15616206.

5. Augustine JJ, Bodziak KA, Hricik DE. Use of sirolimus in solid organ transplantation. Drugs. 2007;67(3):369–91. doi: 10.2165/00003495-200767030-00004. PMID: 17335296.

6. Bell O, Gaberman E, Kilav R, Levi R, Cox KB, Molkentin JD, Silver J, Naveh-Many T. The protein phosphatase calcineurin determines basal parathyroid hormone gene expression. Mol Endocrinol. 2005 Feb;19(2):516–26. doi: 10.1210/me.2004-0108. Epub 2004 Oct 28. PMID: 15514034.

7. Volovelsky O, Cohen G, Kenig A, Wasserman G, Dreazen A, Meyuhas O, Silver J, Naveh-Many T. Phosphorylation of Ribosomal Protein S6 Mediates Mammalian Target of Rapamycin Complex 1-Induced Parathyroid Cell Proliferation in Secondary Hyperparathyroidism. J Am Soc Nephrol. 2016 Apr;27(4):1091–101. doi: 10.1681/ASN.2015040339. Epub 2015 Aug 17. PMID: 26283674; PMCID: PMC4814192.

8. Palchuk MB, London JW, Perez-Rey D, Drebert ZJ, Winer-Jones JP, Thompson CN, Esposito J, Claerhout B. A global federated real-world data and analytics platform for research. JAMIA Open. 2023 May 13;6(2):ooad035. doi: 10.1093/jamiaopen/ooad035. PMID: 37193038; PMCID: PMC10182857.

9. R Core Team (2021). R: A language and environment for statistical computing. R Foundation for Statistical Computing, Vienna, Austria. URL https://www.R-project.org/.

