## Supplementary figures and images for "mTOR Inhibitor-Based Immunosuppression Is Associated with Lower Parathyroid Hormone Levels in Kidney Transplant Recipients: A Multinational Database Analysis and Longitudinal Single-Center Study"

### Supplementary Figure 1

**Supplementary Figure S1. Post-Index Calcium and Phosphate Differences**

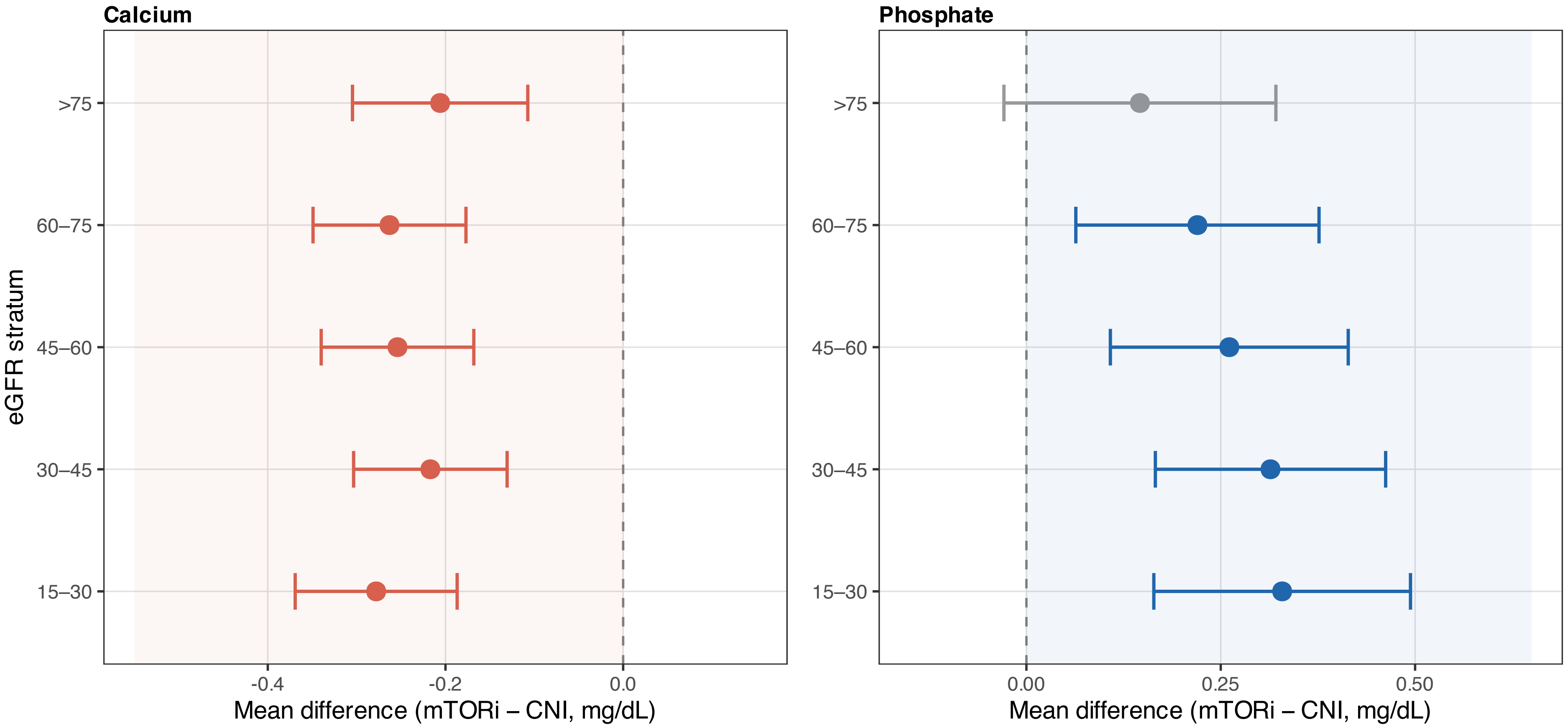
